# DPP6 gene in European American Alzheimer’s Disease

**DOI:** 10.1101/2020.10.23.20216408

**Authors:** Laxmi Kirola, John P. Budde, Fengxian Wang, Joanne Norton, John C. Morris, NIA-LOAD family study group, NCRAD, the ADSP project, Carlos Cruchaga, Maria Victoria Fernández

## Abstract

*DPP6* encodes a transmembrane protein that expresses highly in the hippocampal regions of the brain and regulates dendritic excitability. Recently, rare and loss of function variants were reported in *DPP6* and further demonstrated to be associated with early onset Alzheimer Disease (AD) and frontotemporal dementia. We performed single variant and gene-based analyses in three non-Hispanic white cohorts: a familial late onset AD (cases=1212, controls=341), an unrelated early onset AD (cases=1385, controls=3864) and in the unrelated Alzheimer disease sequencing project (ADSP, cases=5679, controls=4601). Neither single variant or gene-based analysis revealed any significant statistical association of *DPP6* variants with the risk for AD in the cohorts examined.

## 1. Introduction

Alzheimer disease (AD) is the most common neurodegenerative disorder with a complex polygenic inheritance, which is governed by the interplay of aging, environmental, genetic and other risk factors (Bird, 2008; Karch et al., 2014). Previous studies have identified three predominant and highly penetrating Mendelian genes - *APP, PSEN1* and *PSEN2* - and more than 20 loci harboring common and low frequency variants associated with AD (Kunkle et al., 2019). Hence, a large part of AD genetics remains unexplained and demands further investigations to identify novel genes(s) or rare genetic variant(s) associated with AD.

Using linkage analysis in a large Dutch EOAD family, Rademakers et al. (2005) identified a candidate region chromosome 7q36. Follow-up studies of this region performed by Cacace et al., 2019 revealed a chromosomal inversion disrupting the coding sequence of *DPP6* in the Dutch family, as well as several rare non-synonymous variants in a large EOAD Belgian cohort (SKAT-O p-value = 0.03, OR = 2.21, 95% CI 1.05–4.82) (Cacace et al., 2019; Rademakers et al., 2005). *DPP6* encodes a transmembrane protein, predominantly expressed in the brain, that binds to potassium channel Kv4.2 and regulates its gate activity, dendritic excitability and plasticity of hippocampal pyramidal neurons (Lin et al., 2018). Using *in vitro* modeling, Cacace et al. showed that the identified DPP6 missense variants lead to a loss of protein which causes hyperexcitability and behavioral alterations in Dpp6-KO mice. They also found reduced DPP6 and/or Kv4.2 expression in brain tissue of missense variant carriers. No additional studies on this gene have been performed yet; therefore, the contribution of *DPP6* to AD genetics remains unclear. Here, we investigate the potential association of coding variants present in *DPP6* with AD, in three cohorts: the Familial Alzheimer Sequencing (FASe) project, an unrelated early onset AD (EOAD), and the unrelated Alzheimer Disease Sequencing Project (ADSP).

## 2. Methods

We analyzed whole exome and whole genome sequence data from three cohorts: a familial late onset AD (FASe, 525 families, cases=1,212, controls=341) (Fernández et al., 2017, 2016, 2018), an early onset AD (EOAD, cases=1,385, controls=3,864), whose data was generated at Washington University in St. Louis (WASHU) (Fernández et al., 2016, 2018), and a case-control late onset AD (LOAD) from the ADSP (ADSP, cases=5,679, controls=4,601) (Beecham et al., 2017). The ADSP data (pht003392.v7.p4) is available to qualified researchers through the database of Genotypes and Phenotypes (https://www.ncbi.nlm.nih.gov/projects/gap/cgi-bin/dataset.cgi?study_id=phs000572.v8.p4&pht=3392). The WASHU datasets were processed (alignment to GRCh37, variant calling, quality control and annotation) as previously described (Fernández et al., 2017, 2018). Cryptic relatedness (IBD analysis) and population admixture (PCA analysis) were performed in each of the datasets using PLINK 1.9 and only non-Hispanic whites were kept for further analyses (Table 1).

**Table 1.** Demographic characteristics of each of the cohorts employed in this study.

| COHORT | STATUS | N | %Fe | %APOE ε4 | Age (X ± SD) |
| --- | --- | --- | --- | --- | --- |
| FASe | CA | 1,212 | 63.61 | 69.66 | 72.71±9.51 |
|  | CO | 341 | 56.89 | 51.24 | 80.52± 9.70 |
| EOAD | CA | 1,385 | 51.91 | 67.87 | 60.39±2.91 |
|  | CO | 3,864 | 61.05 | 61.05 | 91.27± 8.01 |
| ADSP | CA | 5,656 | 57.00 | 42.35 | 75.50±9.30 |
|  | CO | 4,601 | 59.00 | 14.00 | 87.20± 8.20 |

CA= cases; CO= controls; %Fe= percentage of female; %APOE4= percentage of APOE ε4;

We used the canonical transcript (ENST00000377770.8) of *DPP6* for annotation purposes. Each of the three cohorts was analyzed separately. First, we checked all the reported variants reported in *DPP6* by Cacace et al., 2019. Then, we investigated the effect of *DPP6* variants on the risk of AD by performing single variant logistic regression analysis with sex, PC1 and PC2 as covariates using PLINK 1.9 (Chang et al., 2015) as well as gene burden analysis. We ran two burden tests: (i) non-synonymous rare variants with minor allele frequency-MAF ≤ 1% (MAF≤1%); (ii) non-synonymous variants with a CADD score ≥ 20 (CADD≥20). Gene-based burden analysis was performed in the ADSP and EAOD cohorts using SKAT-O (Wu et al., 2011) after correcting for sex, PC1 and PC2.

## 3. Results

We identified three *DPP6* variants in FASe, eight variants in EOAD and 143 variants in ADSP, including a total of two, seven and 109 coding variants respectively (Supplementary Table 1). A rare synonymous variant (p.Cys735Cys, MAF=3.4-4.9×10^−03^) was found in the three cohorts and seven variants were common between the EOAD and ADSP cohort (7:154237613:A:G splice region, p.Pro249Leu, p.Ser636Cys, p.Thr647Thr, p.Ala655Thr, p.Gln731Gln, p.Leu854Pro). We identified 56 and three rare (MAF<1%) nonsynonymous variants, and 37 and two nonsynonymous variants with a CADD score ≥ 20 in the ADSP and EOAD cohort, respectively. No single variant was significant in any of the cohorts examined (Supplementary Table 1). We could only perform the burden analysis for MAF≤1% or CADD≥20 nonsynonymous variants on the EOAD and ADSP datasets; these were non-significant in the ADSP dataset but showed a trend towards significance in the EOAD dataset (MAF≤1% p-value= 0.061; CADD≥20 p-value = 0.055) (Table2).

**Table 2.** Gene burden analysis on *DPP6* variants in FASe, EOAD and ADSP datasets.

| Cohorts | gene set | N | cummOR | SKAT-O p |
| --- | --- | --- | --- | --- |
| EOAD | MAF≤1% | 3 | 5.32 | 0.061 |
| | CADD≥20 | 2 | NA | 0.055 |
| ADSP | MAF≤1% | 56 | 0.98 | 0.874 |
| | CADD≥20 | 37 | 0.98 | 0.893 |

MAF = minor allele frequency; CADD = combined annotation-dependent depletion; N=number of variants included in the burden analysis.

We crosschecked the variants identified in this study with those reported by Cacace et al. We found that seven missense variants of the 25 reported by Cacace et al. were seen in the ADSP cohort (p.Pro229Thr, p.Arg247His, p.Arg322His, p.His357Arg, p.Lys570Asn, p.Lys571Gln, p.Ala655Thr, p.Ala778Thr) and one of those (p.Ala655Thr) was also present in the EOAD cohort. These variants fall within the extracellular domains (β-propeller and α/β-hydrolase) of *DPP6* protein. Only the p.His357Arg was observed with the same direction of effect (present only in cases) in both Cacace study and the ADSP cohort (Supplementary Table 1).

## 4. Discussion

*DPP6* was recently reported to be associated with EOAD in a Belgian cohort (Cacace et al., 2019). Here, we performed single variant and gene-based association analyses in *DPP6* in three cohorts of non-Hispanic white individuals: FASe (525 families, N=1,553), EOAD (N=5,249) and ADSP (N=10,280). *DPP6* was not found associated with AD risk in none of these cohorts. Neither the recently reported *DPP6* variants by Cacace et al., 2019 nor any other rare variants found in our study seems to be associated to AD, despite our cohorts (FASe, EOAD, and ADSP) were larger than that of Cacace et al., 2019 (CA=221 and CO=237), and we had enough statistical power (96.4%, α=0.05, MAF=0.01, OR=2.00) to replicate their findings.

Cacace et al., 2019 reported a high burden of rare variants in *DPP6* which could be better explained with a possible population isolation effect of *DPP6* variants in Dutch population (Somers et al., 2017). This correlation between rarity of a gene with population specificity has been previously reported for other AD risk loci (Cruchaga et al., 2012; Del-Aguila et al., 2015; Fernández et al., 2016; Wang et al., 2015). Therefore, the lack of association between *DPP6* and AD detected in this study is another example of how population-specific rare variants can be difficult to replicate in other populations, and demonstrates the need to examine independent cohorts before claiming the involvement of a gene with a disease. Accordingly, further populations and ethnicities should be tested for association with DPP6, given that: (i) we found a trend towards association in the EOAD dataset, and (ii) this gene has also been associated with amyotrophic lateral sclerosis (Van Es et al., 2008) mental retardation, autism, muscular dystrophy, and schizophrenia (Liao et al., 2013; Pottier et al., 2019).

## Data Availability

All data used in the manuscript is publicly available at the corresponding repositories (NIAGADs and dbGAP), and available to qualified researchers

## Supplementary Material

**Supplementary Table 1**. Observed DPP6 variants in each of the cohorts investigated: ADSP, EOAD and FASe.

**Supplementary Table 2**. Variants included in the MAF<1% gene-based analyses in each of the cohorts investigated: ADSP, EOAD and FASe.

**Supplementary Table 3**. Variants included in the CADD>20 gene-based analyses in each of the cohorts investigated: ADSP, EOAD and FASe.

## Conflict of interest statement

CC receives research support from: Biogen, EISAI, Alector and Parabon. The funders of the study had no role in the collection, analysis, or interpretation of data; in the writing of the report; or in the decision to submit the paper for publication. CC is a member of the advisory board of Vivid genetics, Halia Therapeutics and ADx Healthcare.

## Acknowledgements

We thank all participants and their families for their commitment and dedication to helping advance research into the early detection and causation of AD. This work was supported by access to the equipment made possible by the Hope Center for Neurological Disorders, and the Departments of Neurology and Psychiatry at Washington University School of Medicine. The authors gratefully acknowledge the study subjects for their participation and the many researchers whose efforts contributed to the assembly and characterization of these datasets.

This work was supported by grants from the NIH to C. Cruchaga (R01AG044546, P01AG003991, RF1AG053303, RF1AG058501, U01AG058922) and to M.V.F (1K99AG061281-01). The recruitment and clinical characterization of research participants at Washington University was supported by NIH P50 AG05681, P01 AG03991, and P01 AG026276.

The Alzheimer’s Disease Sequencing Project (ADSP) is comprised of two Alzheimer’s Disease (AD) genetics consortia and three National Human Genome Research Institute (NHGRI) funded Large Scale Sequencing and Analysis Centers (LSAC). The two AD genetics consortia are the Alzheimer’s Disease Genetics Consortium (ADGC) funded by NIA (U01 AG032984), and the Cohorts for Heart and Aging Research in Genomic Epidemiology (CHARGE) funded by NIA (R01 AG033193), the National Heart, Lung, and Blood Institute (NHLBI), other National Institute of Health (NIH) institutes and other foreign governmental and non-governmental organizations. The Discovery Phase analysis of sequence data is supported through UF1AG047133 (to Drs. Schellenberg, Farrer, Pericak-Vance, Mayeux, and Haines); U01AG049505 to Dr. Seshadri; U01AG049506 to Dr. Boerwinkle; U01AG049507 to Dr. Wijsman; and U01AG049508 to Dr. Goate and the Discovery Extension Phase analysis is supported through U01AG052411 to Dr. Goate, U01AG052410 to Dr. Pericak-Vance and U01 AG052409 to Drs. Seshadri and Fornage. Data generation and harmonization in the Follow-up Phases is supported by U54AG052427 (to Drs. Schellenberg and Wang).

The ADGC cohorts include: Adult Changes in Thought (ACT), the Alzheimer’s Disease Centers (ADC), the Chicago Health and Aging Project (CHAP), the Memory and Aging Project (MAP), Mayo Clinic (MAYO), Mayo Parkinson’s Disease controls, University of Miami, the Multi-Institutional Research in Alzheimer’s Genetic Epidemiology Study (MIRAGE), the National Cell Repository for Alzheimer’s Disease (NCRAD), the National Institute on Aging Late Onset Alzheimer’s Disease Family Study (NIA-LOAD), the Religious Orders Study (ROS), the Texas Alzheimer’s Research and Care Consortium (TARC), Vanderbilt University/Case Western Reserve University (VAN/CWRU), the Washington Heights-Inwood Columbia Aging Project (WHICAP) and the Washington University Sequencing Project (WUSP), the Columbia University Hispanic-Estudio Familiar de Influencia Genetica de Alzheimer (EFIGA), the University of Toronto (UT), and Genetic Differences (GD).

The CHARGE cohorts are supported in part by National Heart, Lung, and Blood Institute (NHLBI) infrastructure grant HL105756 (Psaty), RC2HL102419 (Boerwinkle) and the neurology working group is supported by the National Institute on Aging (NIA) R01 grant AG033193. The CHARGE cohorts participating in the ADSP include the following: Austrian Stroke Prevention Study (ASPS), ASPS-Family study, and the Prospective Dementia Registry-Austria (ASPS/PRODEM-Aus), the Atherosclerosis Risk in Communities (ARIC) Study, the Cardiovascular Health Study (CHS), the Erasmus Rucphen Family Study (ERF), the Framingham Heart Study (FHS), and the Rotterdam Study (RS). ASPS is funded by the Austrian Science Fond (FWF) grant number P20545-P05 and P13180 and the Medical University of Graz. The ASPS-Fam is funded by the Austrian Science Fund (FWF) project I904),the EU Joint Programme - Neurodegenerative Disease Research (JPND) in frame of the BRIDGET project (Austria, Ministry of Science) and the Medical University of Graz and the Steiermärkische Krankenanstalten Gesellschaft. PRODEM-Austria is supported by the Austrian Research Promotion agency (FFG) (Project No. 827462) and by the Austrian National Bank (Anniversary Fund, project 15435. ARIC research is carried out as a collaborative study supported by NHLBI contracts (HHSN268201100005C, HHSN268201100006C, HHSN268201100007C, HHSN268201100008C, HHSN268201100009C, HHSN268201100010C, HHSN268201100011C, and HHSN268201100012C). Neurocognitive data in ARIC is collected by U01 2U01HL096812, 2U01HL096814, 2U01HL096899, 2U01HL096902, 2U01HL096917 from the NIH (NHLBI, NINDS, NIA and NIDCD), and with previous brain MRI examinations funded by R01-HL70825 from the NHLBI. CHS research was supported by contracts HHSN268201200036C, HHSN268200800007C, N01HC55222, N01HC85079, N01HC85080, N01HC85081, N01HC85082, N01HC85083, N01HC85086, and grants U01HL080295 and U01HL130114 from the NHLBI with additional contribution from the National Institute of Neurological Disorders and Stroke (NINDS). Additional support was provided by R01AG023629, R01AG15928, and R01AG20098 from the NIA. FHS research is supported by NHLBI contracts N01-HC-25195 and HHSN268201500001I. This study was also supported by additional grants from the NIA (R01s AG054076, AG049607 and AG033040 and NINDS (R01 NS017950). The ERF study as a part of EUROSPAN (European Special Populations Research Network) was supported by European Commission FP6 STRP grant number 018947 (LSHG-CT-2006-01947) and also received funding from the European Community’s Seventh Framework Programme (FP7/2007-2013)/grant agreement HEALTH-F4-2007-201413 by the European Commission under the programme “Quality of Life and Management of the Living Resources” of 5th Framework Programme (no. QLG2-CT-2002-01254). High-throughput analysis of the ERF data was supported by a joint grant from the Netherlands Organization for Scientific Research and the Russian Foundation for Basic Research (NWO-RFBR 047.017.043). The Rotterdam Study is funded by Erasmus Medical Center and Erasmus University, Rotterdam, the Netherlands Organization for Health Research and Development (ZonMw), the Research Institute for Diseases in the Elderly (RIDE), the Ministry of Education, Culture and Science, the Ministry for Health, Welfare and Sports, the European Commission (DG XII), and the municipality of Rotterdam. Genetic data sets are also supported by the Netherlands Organization of Scientific Research NWO Investments (175.010.2005.011, 911-03-012), the Genetic Laboratory of the Department of Internal Medicine, Erasmus MC, the Research Institute for Diseases in the Elderly (014-93-015; RIDE2), and the Netherlands Genomics Initiative (NGI)/Netherlands Organization for Scientific Research (NWO) Netherlands Consortium for Healthy Aging (NCHA), project 050-060-810. All studies are grateful to their participants, faculty and staff. The content of these manuscripts is solely the responsibility of the authors and does not necessarily represent the official views of the National Institutes of Health or the U.S. Department of Health and Human Services.

The four LSACs are: the Human Genome Sequencing Center at the Baylor College of Medicine (U54 HG003273), the Broad Institute Genome Center (U54HG003067), The American Genome Center at the Uniformed Services University of the Health Sciences (U01AG057659), and the Washington University Genome Institute (U54HG003079).

Biological samples and associated phenotypic data used in primary data analyses were stored at Study Investigators institutions, and at the National Cell Repository for Alzheimer’s Disease (NCRAD, U24AG021886) at Indiana University funded by NIA. Associated Phenotypic Data used in primary and secondary data analyses were provided by Study Investigators, the NIA funded Alzheimer’s Disease Centers (ADCs), and the National Alzheimer’s Coordinating Center (NACC, U01AG016976) and the National Institute on Aging Genetics of Alzheimer’s Disease Data Storage Site (NIAGADS, U24AG041689) at the University of Pennsylvania, funded by NIA, and at the Database for Genotypes and Phenotypes (dbGaP) funded by NIH. This research was supported in part by the Intramural Research Program of the National Institutes of health, National Library of Medicine. Contributors to the Genetic Analysis Data included Study Investigators on projects that were individually funded by NIA, and other NIH institutes, and by private U.S. organizations, or foreign governmental or nongovernmental organizations.

See Supplementary Material

## Notes

### Competing Interest Statement

The authors have declared no competing interest.

### Author Declarations

The approval number for the Knight ADRC Genetics Core family studies is 201104178. For the EOAD samples the PMID is 30739198. We received dbGAP approval to use ADSP data; our approval number is #6109, under the project title: "Identification of novel genetic variants and genes implicated on neurodegenerative diseases and other and comorbid psychiatric disorders"; the IRB for the previous project is #: 201304112.

## References

Beecham, G.W., Bis, J.C., Martin, E.R., Choi, S.H., DeStefano, A.L., Van Duijn, C.M., Fornage, M., Gabriel, S.B., Koboldt, D.C., Larson, D.E., Naj, A.C., Psaty, B.M., Salerno, W., Bush, W.S., Foroud, T.M., Wijsman, E., Farrer, L.A., Goate, A., Haines, J.L., Pericak-Vance, M.A., Boerwinkle, E., Mayeux, R., Seshadri, S., Schellenberg, G., 2017. Clinical/Scientific Notes: The Alzheimer’s disease sequencing project: Study design and sample selection. Neurol. Genet. https://doi.org/10.1212/NXG.0000000000000194

Bird, T.D., 2008. Genetic aspects of Alzheimer disease. Genet. Med. https://doi.org/10.1097/GIM.0b013e31816b64dc

Cacace, R., Heeman, B., Van Mossevelde, S., De Roeck, A., Hoogmartens, J., De Rijk, P., Gossye, H., De Vos, K., De Coster, W., Strazisar, M., De Baets, G., Schymkowitz, J., Rousseau, F., Geerts, N., De Pooter, T., Peeters, K., Sieben, A., Martin, J.J., Engelborghs, S., Salmon, E., Santens, P., Vandenberghe, R., Cras, P., P. De Deyn, P. C. van Swieten, J. M. van Duijn C., van der Zee, J., Sleegers, K., Van Broeckhoven, C., Goeman, J., Crols, R., Nuytten, D., De Bleecker, J.L., Van Langenhove, T., Ivanoiu, A., Deryck, O., Bergmans, B., Versijpt, J., Michotte, A., Delbeck, J., Willems, C., De Klippel, N., 2019. Loss of DPP6 in neurodegenerative dementia: a genetic player in the dysfunction of neuronal excitability. Acta Neuropathol. https://doi.org/10.1007/s00401-019-01976-3

Chang, C.C., Chow, C.C., Tellier, L.C., Vattikuti, S., Purcell, S.M., Lee, J.J., 2015. Second-generation PLINK: rising to the challenge of larger and richer datasets. Gigascience 4, 7. https://doi.org/10.1186/s13742-015-0047-8

Cruchaga, C., Haller, G., Chakraverty, S., Mayo, K., Vallania, F.L.M., Mitra, R.D., Faber, K., Williamson, J., Bird, T., Diaz-Arrastia, R., Foroud, T.M., Boeve, B.F., Graff-Radford, N.R., St Jean, P., Lawson, M., Ehm, M.G., Mayeux, R., Goate, A.M., 2012. Rare variants in APP, PSEN1 and PSEN2 increase risk for AD in late-onset Alzheimer’s disease families. PLoS One 7, e31039. https://doi.org/10.1371/journal.pone.0031039

Del-Aguila, J.L., Fernández, M.V., Jimenez, J., Black, K., Ma, S., Deming, Y., Carrell, D., Saef, B., Alzheimer’s Disease Neuroimaging Initiative, Howells, B., Budde, J., Cruchaga, C., 2015. Role of ABCA7 loss-of-function variant in Alzheimer’s disease: a replication study in European-Americans. Alzheimers. Res. Ther. 7, 73. https://doi.org/10.1186/s13195-015-0154-x

Del-Aguila, J.L., Fernández, M.V., Schindler, S., Ibanez, L., Deming, Y., Ma, S., Saef, B., Black, K., Budde, J., Norton, J., Chasse, R., Harari, O., Goate, A., Xiong, C., Morris, J.C., Cruchaga, C., Cruchaga, C., 2018. Assessment of the Genetic Architecture of Alzheimer’s Disease Risk in Rate of Memory Decline. J. Alzheimer’s Dis. 62, 745–756. https://doi.org/10.3233/JAD-170834

Fernández, M.V., Black, K., Carrell, D., Saef, B., Budde, J., Deming, Y., Howells, B., Del-Aguila, J.L., Ma, S., Bi, C., Norton, J., Chasse, R., Morris, J., Goate, A., Cruchaga, C.,NIA-LOAD family study group, NCRAD, 2016. SORL1 variants across Alzheimer’s disease European American cohorts. Eur. J. Hum. Genet. https://doi.org/10.1038/ejhg.2016.122

Fernández, M.V., Kim, J.H., Budde, J.P., Black, K., Medvedeva, A., Saef, B., Deming, Y., Del-Aguila, J., Ibañez, L., Dube, U., Harari, O., Norton, J., Chasse, R., Morris, J.C., Goate, A., Cruchaga, C., Ncrad, Cruchaga C., 2017. Analysis of neurodegenerative Mendelian genes in clinically diagnosed Alzheimer Disease. PLOS Genet. 13, e1007045. https://doi.org/10.1371/journal.pgen.1007045

Fernández, M. V., Budde, J., Del-Aguila, J.L., Ibañez, L., Deming, Y., Harari, O., Norton, J., Morris, J.C., Goate, A.M., Cruchaga, C., Mayeux, R., Farlow, M., Foroud, T., Faber, K., Boeve, B.F., Graff-Radford, N.R., Bennett, D.A., Sweet, R.A., Rosenberg, R., Bird, T.D., Silverman, J.M., 2018. Evaluation of gene-based family-based methods to detect novel genes associated with familial late onset Alzheimer disease. Front. Neurosci. 12. https://doi.org/10.3389/fnins.2018.00209

Karch, C.M., Cruchaga, C., Goate, A.M., 2014. Alzheimer’s disease genetics: From the bench to the clinic. Neuron 83, 11–26. https://doi.org/10.1016/j.neuron.2014.05.041

Kunkle, B.W., Grenier-Boley, B., Sims, R., Bis, J.C., Damotte, V., Naj, A.C., Boland, A., Vronskaya, M., van der Lee, S.J., Amlie-Wolf, A., Bellenguez, C., Frizatti, A., Chouraki, V., Martin, E.R., Sleegers, K., Badarinarayan, N., Jakobsdottir, J., Hamilton-Nelson, K.L., Moreno-Grau, S., Olaso, R., Raybould, R., Chen, Y., Kuzma, A.B., Hiltunen, M., Morgan, T., Ahmad, S., Vardarajan, B.N., Epelbaum, J., Hoffmann, P., Boada, M., Beecham, G.W., Garnier, J.-G., Harold, D., Fitzpatrick, A.L., Valladares, O., Moutet, M.-L., Gerrish, A.,Smith, A. V., Qu, L., Bacq, D., Denning, N., Jian, X., Zhao, Y., Del Zompo, M., Fox, N.C., Choi, S.-H., Mateo, I., Hughes, J.T., Adams, H.H., Malamon, J., Sanchez-Garcia, F., Patel, Y., Brody, J.A., Dombroski, B.A., Naranjo, M.C.D., Daniilidou, M., Eiriksdottir, G., Mukherjee, S., Wallon, D., Uphill, J., Aspelund, T., Cantwell, L.B., Garzia, F., Galimberti, D., Hofer, E., Butkiewicz, M., Fin, B., Scarpini, E., Sarnowski, C., Bush, W.S., Meslage, S., Kornhuber, J., White, C.C., Song, Y., Barber, R.C., Engelborghs, S., Sordon, S., Voijnovic, D., Adams, P.M., Vandenberghe, R., Mayhaus, M., Cupples, L.A., Albert, M.S., De Deyn, P.P., Gu, W., Himali, J.J., Beekly, D., Squassina, A., Hartmann, A.M., Orellana, A., Blacker, D., Rodriguez-Rodriguez, E., Lovestone, S., Garcia, M.E., Doody, R.S., Munoz-Fernadez, C., Sussams, R., Lin, H., Fairchild, T.J., Benito, Y.A., Holmes, C., Karamujić-Čomić, H., Frosch, M.P., Thonberg, H., Maier, W., Roschupkin, G., Ghetti, B., Giedraitis, V., Kawalia, A., Li, S., Huebinger, R.M., Kilander, L., Moebus, S., Hernández, I., Kamboh, M.I., Brundin, R., Turton, J., Yang, Q., Katz, M.J., Concari, L., Lord, J., Beiser, A.S., Keene, C.D., Helisalmi, S., Kloszewska, I., Kukull, W.A., Koivisto, A.M., Lynch, A., Tarraga, L., Larson, E.B., Haapasalo, A., Lawlor, B., Mosley, T.H., Lipton, R.B., Solfrizzi, V., Gill, M., Longstreth, W.T., Montine, T.J., Frisardi, V., Diez-Fairen, M., Rivadeneira, F., Petersen, R.C., Deramecourt, V., Alvarez, I., Salani, F., Ciaramella, A., Boerwinkle, E., Reiman, E.M., Fievet, N., Rotter, J.I., Reisch, J.S., Hanon, O., Cupidi, C., Andre Uitterlinden, A.G., Royall, D.R., Dufouil, C., Maletta, R.G., de Rojas, I., Sano, M., Brice, A., Cecchetti, R., George-Hyslop, P.S., Ritchie, K., Tsolaki, M., Tsuang, D.W., Dubois, B., Craig, D., Wu, C.-K., Soininen, H., Avramidou, D., Albin, R.L., Fratiglioni, L., Germanou, A., Apostolova, L.G., Keller, L., Koutroumani, M., Arnold, S.E., Panza, F., Gkatzima, O., Asthana, S., Hannequin, D., Whitehead, P., Atwood, C.S., Caffarra, P., Hampel, H., Quintela, I., Carracedo, Á., Lannfelt, L., Rubinsztein, D.C., Barnes, L.L., Pasquier, F., Frölich, L., Barral, S., McGuinness, B., Beach, T.G., Johnston, J.A., Becker, J.T., Passmore, P., Bigio, E.H., Schott, J.M., Bird, T.D., Warren, J.D., Boeve, B.F., Lupton, M.K., Bowen, J.D., Proitsi, P., Boxer, A., Powell, J.F., Burke, J.R., Kauwe, J.S.K., Burns, J.M., Mancuso, M., Buxbaum, J.D., Bonuccelli, U., Cairns, N.J., McQuillin, A., Cao, C., Livingston, G., Carlson, C.S., Bass, N.J., Carlsson, C.M., Hardy, J., Carney, R.M., Bras, J., Carrasquillo, M.M., Guerreiro, R., Allen, M., Chui, H.C., Fisher, E., Masullo, C., Crocco, E.A., DeCarli, C., Bisceglio, G., Dick, M., Ma, L., Duara, R., Graff-Radford, N.R., Evans, D.A., Hodges, A., Faber, K.M., Scherer, M., Fallon, K.B., Riemenschneider, M., Fardo, D.W., Heun, R., Farlow, M.R., Kölsch, H., Ferris, S., Leber, M., Foroud, T.M., Heuser, I., Galasko, D.R., Giegling, I., Gearing, M., Hüll, M., Geschwind, D.H., Gilbert, J.R., Morris, J., Green, R.C., Mayo, K., Growdon, J.H., Feulner, T., Hamilton, R.L., Harrell, L.E., Drichel, D., Honig, L.S., Cushion, T.D., Huentelman, M.J., Hollingworth, P., Hulette, C.M., Hyman, B.T., Marshall, R., Jarvik, G.P., Meggy, A., Abner, E., Menzies, G.E., Jin, L.-W., Leonenko, G., Real, L.M., Jun, G.R., Baldwin, C.T., Grozeva, D., Karydas, A., Russo, G., Kaye, J.A., Kim, R., Jessen, F., Kowall, N.W., Vellas, B., Kramer, J.H., Vardy, E., LaFerla, F.M., Jöckel, K.-H., Lah, J.J., Dichgans, M., Leverenz, J.B., Mann, D., Levey, A.I., Pickering-Brown, S., Lieberman, A.P., Klopp, N., Lunetta, K.L., Wichmann, H.-E., Lyketsos, C.G., Morgan, K., Marson, D.C., Brown, K., Martiniuk, F., Medway, C., Mash, D.C., Nöthen, M.M., Masliah, E., Hooper, N.M., McCormick, W.C., Daniele, A., McCurry, S.M., Bayer, A., McDavid, A.N., Gallacher, J., McKee, A.C., van den Bussche, H., Mesulam, M., Brayne, C., Miller, B.L., Riedel-Heller, S., Miller, C.A., Miller, J.W., Al-Chalabi, A., Morris, J.C., Shaw, C.E., Myers, A.J., Wiltfang, J., O’Bryant, S., Olichney, J.M., Alvarez, V., Parisi, J.E., Singleton, A.B., Paulson, H.L., Collinge, J., Perry, W.R., Mead, S., Peskind, E., Cribbs, D.H., Rossor, M., Pierce, A., Ryan, N.S., Poon, W.W., Nacmias, B., Potter, H., Sorbi, S., Quinn, J.F., Sacchinelli, E., Raj, A., Spalletta, G., Raskind, M., Caltagirone, C., Bossù, P., Orfei, M.D., Reisberg, B., Clarke, R., Reitz, C., Smith, A.D., Ringman, J.M., Warden, D., Roberson, E.D., Wilcock, G., Rogaeva, E., Bruni, A.C., Rosen, H.J., Gallo, M., Rosenberg, R.N., Ben-Shlomo, Y., Sager, M.A., Mecocci, P., Saykin, A.J., Pastor, P., Cuccaro, M.L., Vance, J.M., Schneider, J.A., Schneider, L.S., Slifer, S., Seeley, W.W., Smith, A.G., Sonnen, J.A., Spina, S., Stern, R.A., Swerdlow, R.H., Tang, M., Tanzi, R.E., Trojanowski, J.Q., Troncoso, J.C., Van Deerlin, V.M., Van Eldik, L.J., Vinters, H. V., Vonsattel, J.P., Weintraub, S., Welsh-Bohmer, K.A., Wilhelmsen, K.C., Williamson, J., Wingo, T.S., Woltjer, R.L., Wright, C.B., Yu, C.-E., Yu, L., Saba, Y., Pilotto, A., Bullido, M.J., Peters, O., Crane, P.K., Bennett, D., Bosco, P., Coto, E., Boccardi, V., De Jager, P.L., Lleo, A., Warner, N., Lopez, O.L., Ingelsson, M., Deloukas, P., Cruchaga, C., Graff, C., Gwilliam, R., Fornage, M., Goate, A.M., Sanchez-Juan, P., Kehoe, P.G., Amin, N., Ertekin-Taner, N., Berr, C., Debette, S., Love, S., Launer, L.J., Younkin, S.G., Dartigues, J.-F., Corcoran, C., Ikram, M.A., Dickson, D.W., Nicolas, G., Campion, D., Tschanz, J., Schmidt, H., Hakonarson, H., Clarimon, J., Munger, R., Schmidt, R., Farrer, L.A., Van Broeckhoven, C. C. O’Donovan, M., DeStefano, A.L., Jones, L., Haines, J.L., Deleuze, J.-F., Owen, M.J., Gudnason, V., Mayeux, R., Escott-Price, V., Psaty, B.M., Ramirez, A., Wang, L.-S., Ruiz, A., van Duijn, C.M., Holmans, P.A., Seshadri, S., Williams, J., Amouyel, P., Schellenberg, G.D., Lambert, J.-C., Pericak-Vance, M.A., 2019. Genetic meta-analysis of diagnosed Alzheimer’s disease identifies new risk loci and implicates Aβ, tau, immunity and lipid processing. Nat. Genet. 51, 414–430. https://doi.org/10.1038/s41588-019-0358-2

Liao, C., Fu, F., Li, R. Yang, W. qing, Liao, H. yi, Yan, J. rong, Li, J., Li, S. yuan, Yang X., Li, D. zhi, 2013. Loss-of-function variation in the DPP6 gene is associated with autosomal dominant microcephaly and mental retardation. Eur. J. Med. Genet. https://doi.org/10.1016/j.ejmg.2013.06.008

Lin, L., Murphy, J.G., Karlsson, R.M., Petralia, R.S., Gutzmann, J.J., Abebe, D., Wang, Y.X., Cameron, H.A., Hoffman, D.A., 2018. DPP6 loss impacts hippocampal synaptic development and induces behavioral impairments in recognition, learning and memory. Front. Cell. Neurosci. https://doi.org/10.3389/fncel.2018.00084

Pottier, C., Ren, Y., Perkerson, R.B., Baker, M., Jenkins, G.D., van Blitterswijk, M., DeJesus-Hernandez, M., van Rooij, J.G.J., Murray, M.E., Christopher, E., McDonnell, S.K., Fogarty, Z., Batzler, A., Tian, S., Vicente, C.T., Matchett, B., Karydas, A.M., Hsiung, G.Y.R., Seelaar, H., Mol, M.O., Finger, E.C., Graff, C., Öijerstedt, L., Neumann, M., Heutink, P., Synofzik, M., Wilke, C., Prudlo, J., Rizzu, P., Simon-Sanchez, J., Edbauer, D., Roeber, S., Diehl-Schmid, J., Evers, B.M., King, A., Mesulam, M.M., Weintraub, S., Geula, C., Bieniek, K.F., Petrucelli, L., Ahern, G.L., Reiman, E.M., Woodruff, B.K., Caselli, R.J., Huey, E.D., Farlow, M.R., Grafman, J., Mead, S., Grinberg, L.T., Spina, S., Grossman, M., Irwin, D.J., Lee, E.B., Suh, E.R., Snowden, J., Mann, D., Ertekin-Taner, N., Uitti, R.J., Wszolek, Z.K., Josephs, K.A., Parisi, J.E., Knopman, D.S., Petersen, R.C., Hodges, J.R., Piguet, O., Geier, E.G., Yokoyama, J.S., Rissman, R.A., Rogaeva, E., Keith, J., Zinman, L., Tartaglia, M.C., Cairns, N.J., Cruchaga, C., Ghetti, B., Kofler, J., Lopez, O.L., Beach, T.G., Arzberger, T., Herms, J., Honig, L.S., Vonsattel, J.P., Halliday, G.M., Kwok, J.B., White, C.L., Gearing, M., Glass, J., Rollinson, S., Pickering-Brown, S., Rohrer, J.D., Trojanowski, J.Q., Van Deerlin, V., Bigio, E.H., Troakes, C., Al-Sarraj, S., Asmann, Y., Miller, B.L., Graff-Radford, N.R., Boeve, B.F., Seeley, W.W., Mackenzie, I.R.A., van Swieten, J.C., Dickson, D.W., Biernacka, J.M., Rademakers, R., 2019. Genome-wide analyses as part of the international FTLD-TDP whole-genome sequencing consortium reveals novel disease risk factors and increases support for immune dysfunction in FTLD. Acta Neuropathol. https://doi.org/10.1007/s00401-019-01962-9

Rademakers, R., Cruts, M., Sleegers, K., Dermaut, B., Theuns, J., Aulchenko, Y., Weckx, S., De Pooter, T., Van Den Broeck, M., Corsmit, E., De Rijk, P., Del-Favero, J., Van Swieten, J., Van Duijn, C.M., Van Broeckhoven, C., 2005. Linkage and association studies identify a novel locus for Alzheimer disease at 7q36 in a Dutch population-based sample. Am. J. Hum. Genet. https://doi.org/10.1086/491749

Somers, M., Olde Loohuis, L.M., Aukes, M.F., Pasaniuc, B., De Visser, K.C.L., Kahn, R.S., Sommer, I.E., Ophoff, R.A., 2017. A genetic population isolate in the Netherlands showing extensive haplotype sharing and long regions of homozygosity. Genes (Basel). https://doi.org/10.3390/genes8050133

Van Es, M.A., Van Vught, P.W.J., Blauw, H.M., Franke, L., Saris, C.G.J., Van Den Bosch, L., De Jong, S.W., De Jong, V., Baas, F., Van’t Slot, R., Lemmens, R., Schelhaas, H.J., Birve, A., Sleegers, K., Van Broeckhoven, C., Schymick, J.C., Traynor, B.J., Wokke, J.H.J., Wijmenga, C., Robberecht, W., Andersen, P.M., Veldink, J.H., Ophoff, R.A., Van Den Berg, L.H., 2008. Genetic variation in DPP6 is associated with susceptibility to amyotrophic lateral sclerosis. Nat. Genet. https://doi.org/10.1038/ng.2007.52

Wang, L.S., Naj, A.C., Graham, R.R., Crane, P.K., Kunkle, B.W., Cruchaga, C., Gonzalez Murcia, J.D., Cannon-Albright, L., Baldwin, C.T., Zetterberg, H., Blennow, K., Kukull, W.A., Faber, K.M., Schupf, N., Norton, M.C., Tschanz, J.A.T., Munger, R.G., Corcoran, C.D., Rogaeva, E., Albert, M.S., Albin, R.L., Apostolova, L.G., Arnold, S.E., Barber, R., Barmada, M.M., Barnes, L.L., Beach, T.G., Becker, J.T., Beecham, G.W., Beekly, D., Bennett, D.A., Bigio, E.H., Bird, T.D., Blacker, D., Boeve, B.F., Bowen, J.D., Boxer, A., Burke, J.R., Buxbaum, J.D., Cairns, N.J., Cao, C., Carlson, C.S., Carroll, S.L., Chui, H.C., Clark, D.G., Cribbs, D.H., Crocco, E.A., DeCarli, C., DeKosky, S.T., Demirci, F.Y., Dick, M., Dickson, D.W., Duara, R., Ertekin-Taner, N., Fallon, K.B., Farlow, M.R., Ferris, S., Frosch, M.P., Galasko, D.R., Ganguli, M., Gearing, M., Geschwind, D.H., Ghetti, B., Gilbert, J.R., Glass, J.D., Graff-Radford, N.R., Growdon, J.H., Hamilton, R.L., Hamilton-Nelson, K.L., Harrell, L.E., Head, E., Honig, L.S., Hulette, C.M., Hyman, B.T., Jarvik, G.P., Jicha, G.A., Jin, L.W., Jun, G., Kamboh, M.I., Karydas, A., Kaye, J.A., Kim, R., Koo, E.H., Kowall, N.W., Kramer, J.H., Kramer, P., LaFerla, F.M., Lah, J.J., Leverenz, J.B., Levey, A.I., Li, G., Lieberman, A.P., Lopez, O.L., Lunetta, K.L., Lyketsos, C.G., Mack, W.J., Marson, D.C., Martin, E.R., Martiniuk, F., Mash, D.C., Masliah, E., McCormick, W.C., McCurry, S.M., McDavid, A.N., McKee, A.C., Mesulam, M.M., Miller, B.L., Miller, C.A., Miller, J.W., Montine, T.J., Morris, J.C., Murrell, J.R., Olichney, J.M., Parisi, J.E., Perry, W., Peskind, E., Petersen, R.C., Pierce, A., Poon, W.W., Potter, H., Quinn, J.F., Raj, A., Raskind, M., Reiman, E.M., Reisberg, B., Reitz, C., Ringman, J.M., Roberson, E.D., Rosen, H.J., Rosenberg, R.N., Sano, M., Saykin, A.J., Schneider, J.A., Schneider, L.S., Seeley, W.W., Smith, A.G., Sonnen, J.A., Spina, S., Stern, R.A., Tanzi, R.E., Thornton-Wells, T.A., Trojanowski, J.Q., Troncoso, J.C., Tsuang, D.W., Van Deerlin, V.M., Van Eldik, L.J., Vardarajan, B.N., Vinters, H. V., Vonsattel, J.P., Weintraub, S., Welsh-Bohmer, K.A., Williamson, J., Wishnek, S., Woltjer, R.L., Wright, C.B., Younkin, S.G., Yu, C.E., Yu, L., Lin, C.F., Dombroski, B.A., Cantwell, L.B., Partch, A., Valladares, O., Hakonarson, H., St George-Hyslop, P., Green, R.C., Goate, A.M., Foroud, T.M., Carney, R.M., Larson, E.B., Behrens, T.W., Kauwe, J.S.K., Haines, J.L., Farrer, L.A., Pericak-Vance, M.A., Mayeux, R., Schellenberg, G.D., 2015. Rarity of the alzheimer disease-protective APP A673T variant in the United States. JAMA Neurol. https://doi.org/10.1001/jamaneurol.2014.2157

Wu, M.C., Lee, S., Cai, T., Li, Y., Boehnke, M., Lin, X., 2011. Rare-variant association testing for sequencing data with the sequence kernel association test. Am. J. Hum. Genet. 89, 82–93. https://doi.org/10.1016/j.ajhg.2011.05.029

