## Supplementary-table1 for "DPP6 gene in European American Alzheimer’s Disease": 2020-10-DPP6-Sup-table-1.pdf

**Supplementary Table 1.** Observed *DPP6* variants in each of the cohorts investigated: ADSP, EOAD and FASe.

| SNP-ID* | EFFECT | p.CHAN GE | direction in Cacace | CADD | MAF-ExAC** | MAF | Cases (n=5656) |
|---|---|---|---|---|---|---|---|
| 7:154002603:C:T | intron variant | . | . | . | . | 4.89E-05 | 0 |
| 7:154002619:G:A | intron variant | . | . | 20.6 | 8.31E-06 | 4.91E-05 | 0 |
| 7:154143322:G:A | synonymous variant | p.Pro89Pro | . | . | . | . | . |
| 7:154143346:A:G | synonymous variant | p.Ala97Ala | . | . | . | 4.86E-05 | 1 |
| 7:154172020:C:G | splice region variant&intron variant | . | . | . | . | 4.87E-05 | 1 |
| 7:154172040:G:A | synonymous variant | p.Leu125Leu | . | . | . | 4.86E-05 | 1 |
| 7:154172046:A:G | synonymous variant | p.Gln127Gln | . | . | . | 9.73E-05 | 1 |
| 7:154172103:C:T | synonymous variant | p.Pro146Pro | . | . | . | 9.73E-05 | 1 |
| 7:154237613:A:G | splice region variant&intron variant | . | . | . | . | 9.25E-04 | 10 |
| 7:154237630:C:T | synonymous variant | p.Ile157Ile | . | . | . | 4.87E-05 | 0 |
| 7:154237645:A:G | synonymous variant | p.Lys162Lys | . | . | . | 4.87E-05 | 1 |
| 7:154237680:C:T | missense variant | p.Thr174Ile | . | 0.002 | . | 4.87E-05 | 1 |
| 7:154263937:G:A | missense variant | p.Arg188Lys | . | 12.83 | . | 4.87E-05 | 1 |
| 7:154263950:T:C | synonymous variant | p.Tyr192Tyr | . | . | . | 3.89E-04 | 4 |
| 7:154263951:G:A | missense variant | p.Glu193Lys | . | 22.8 | 2.48E-05 | 4.87E-05 | 1 |
| 7:154263988:A:G | missense variant | p.Tyr205Cys | . | 25.8 | . | 4.87E-05 | 0 |
| 7:154379370:T:A | intron variant | . | . | 10.69 | . | 4.87E-05 | 0 |
| 7:154379448:G:C | intron variant | . | . | 7.985 | . | 4.86E-05 | 0 |
| 7:154379453:C:T | intron variant | . | . | 9.289 | . | 1.46E-04 | 0 |
| 7:154379480:A:G | intron variant | . | . | 5.343 | . | 9.73E-05 | 1 |
| 7:154379487:T:G | intron variant | . | . | 8.74 | 8.60E-06 | 4.86E-05 | 0 |
| 7:154379517:C:T | intron variant | . | . | 5.269 | 8.85E-02 | 9.62E-02 | 56/1003/4616 |
| 7:154379570:C:T | intron variant | . | . | 5.738 | 1.10E-02 | 1.54E-02 | 2/177/5498 |
| 7:154379583:T:C | intron variant | . | . | 1.54 | 8.57E-05 | 4.86E-05 | 1 |
| 7:154379634:C:A | intron variant | . | . | 6.007 | 8.55E-06 | 4.87E-05 | 0 |
| 7:154379645:A:G | intron variant | . | . | 2.193 | . | 4.87E-05 | 1 |
| 7:154379646:C:T | intron variant | . | . | 3.689 | 5.91E-04 | 1.46E-04 | 1 |
| 7:154379655:G:A | intron variant | . | . | 2.92 | 7.70E-05 | 4.87E-05 | 0 |
| 7:154379688:G:A | intron variant | . | . | 1.987 | 1.20E-04 | 1.46E-04 | 0 |
| 7:154379701:C:T | intron variant | . | . | . | . | 9.76E-05 | 1 |

| | | | | | | | |
|---|---|---|---|---|---|---|---|
| 7:154379718:A:T | intron variant | . | . | 3.409 | 2.57E-05 | 4.88E-05 | 1 |
| 7:154379727:A:G | intron variant | . | . | 6.989 | 3.14E-03 | 5.80E-03 | 67 |
| 7:154379739:C:T | intron variant | . | . | 1.279 | 8.98E-02 | 7.91E-02 | 50/731/4537 |
| 7:154379742:G:T | intron variant | . | . | 1.272 | 1.09E-02 | 1.55E-02 | 2/175/5483 |
| 7:154379799:T:C | intron variant | . | . | . | . | 5.02E-05 | 0 |
| 7:154379800:G:A | intron variant | . | . | . | . | 1.01E-04 | 2 |
| 7:154429560:C:T | synonymous variant | p.Tyr219Tyr | . | . | . | 2.87E-01 | 491/2287/2891 |
| 7:154429569:C:T | synonymous variant | p.Ser222Ser | . | . | . | 5.45E-03 | 1/65/5612 |
| 7:154429580:A:G | missense variant | p.His226Arg | . | 21.2 | . | 4.87E-05 | 1 |
| ***7:154461074:C:A*** | ***missense variant*** | ***p.Pro229Thr*** | ***CA*** | ***22.1*** | ***.*** | ***1.95E-04*** | ***3*** |
| 7:154461105:A:C | missense variant | p.Asn239Thr | . | 23.4 | . | 4.86E-05 | 1 |
| 7:154461112:A:G | synonymous variant | p.Lys241Lys | . | . | . | 9.65E-02 | 56/991/4632 |
| 7:154461215:A:G | intron variant | . | . | . | . | . | . |
| 7:154461135:C:T | missense variant | p.Pro249Leu | . | 27.3 | . | 4.86E-05 | 1 |
| 7:154461145:A:G | synonymous variant | p.Gln252Gln | . | . | . | 4.86E-05 | 1 |
| 7:154519513:C:T | missense variant | p.His267Tyr | . | 23.8 | 3.31E-05 | 4.86E-05 | 0 |
| 7:154519527:G:A | synonymous variant | p.Gln271Gln | . | . | . | 6.32E-04 | 8 |
| ***7:154519535:G:A*** | ***missense variant*** | ***p.Arg274His*** | ***CA*** | ***34*** | ***8.27E-06*** | ***4.86E-05*** | ***0*** |
| 7:154519551:C:T | synonymous variant | p.Gly279Gly | . | . | . | 4.86E-05 | 1 |
| 7:154519584:C:T | synonymous variant | p.Asp290Asp | . | . | . | 4.86E-05 | 0 |
| 7:154519593:T:C | synonymous variant | p.Tyr293Tyr | . | . | . | 4.86E-05 | 0 |
| 7:154561149:C:T | synonymous variant | p.Ile302Ile | . | . | . | 4.86E-05 | 1 |
| 7:154561167:G:A | synonymous variant | p.Pro308Pro | . | . | . | 1.95E-04 | 3 |
| 7:154561176:G:A | synonymous variant | p.Thr311Thr | . | . | . | 9.73E-05 | 1 |
| 7:154561182:C:T | synonymous variant | p.Leu313Leu | . | . | . | 4.86E-05 | 1 |
| 7:154561183:G:A | missense variant | p.Ala314Thr | . | 27.1 | 3.31E-05 | 4.86E-05 | 1 |
| 7:154561188:C:T | synonymous variant | p.Tyr315Tyr | . | . | . | 8.57E-02 | 50/865/4762 |
| ***7:154561208:G:A*** | ***missense variant*** | ***p.Arg322His*** | ***CA*** | ***25.8*** | ***1.08E-04*** | ***9.73E-05*** | ***1*** |
| 7:154561218:C:G | missense variant | p.Ile325Met | . | 11.86 | 9.10E-05 | 4.86E-05 | 0 |
| 7:154561240:G:A | missense variant | p.Gly333Ser | . | 25 | 1.66E-04 | 4.87E-05 | 1 |
| 7:154561254:C:G | synonymous variant | p.Pro337Pro | . | . | . | 4.88E-05 | 0 |
| 7:154561276:C:T | missense variant | p.Pro345Ser | . | 28.5 | 8.27E-06 | 4.94E-05 | 1 |
| 7:154564555:G:T | missense variant&splice region variant | p.Ala347Ser | . | 26 | 8.27E-06 | 4.86E-05 | 1 |
| ***7:154564586:A:G*** | ***missense variant*** | ***p.His357Arg*** | ***CA*** | ***21.7*** | ***.*** | ***9.73E-05*** | ***2*** |

| | | | | | | | |
|---|---|---|---|---|---|---|---|
| 7:154564591:A:G | missense variant | p.Ile359Val | . | 11.25 | . | 4.86E-05 | 0 |
| 7:154564605:A:T | synonymous variant | p.Gly363Gly | . | . | . | 4.86E-05 | 1 |
| 7:154564646:G:T | missense variant | p.Arg377Leu | . | 27.6 | . | 4.88E-05 | 1 |
| 7:154585780:C:T | intron variant | . | . | . | . | 4.87E-05 | 0 |
| 7:154585806:T:C | missense variant | p.Met385Thr | . | 18.77 | 3.30E-05 | 4.86E-05 | 1 |
| 7:154587563:G:T | missense variant | p.Glu423Asp | . | 25.2 | . | 4.86E-05 | 0 |
| 7:154587577:C:T | missense variant | p.Ala428Val | . | 25.1 | . | 4.86E-05 | 1 |
| 7:154587586:AC:A | frameshift variant | p.His431fs | . | . | . | 4.87E-05 | 1 |
| 7:154593078:T:C | missense variant | p.Val438Ala | . | 22.9 | . | 4.86E-05 | 0 |
| 7:154593084:C:T | missense variant | p.Ser440Phe | . | 34 | . | 4.86E-05 | 0 |
| 7:154593156:C:T | missense variant | p.Thr464Met | . | 23 | . | 4.88E-05 | 1 |
| 7:154593157:G:A | synonymous variant | p.Thr464Thr | . | . | . | 1.46E-04 | 0 |
| 7:154595580:A:G | missense variant | p.Ser472Gly | . | 22.3 | . | 4.86E-05 | 1 |
| 7:154595624:C:T | synonymous variant | p.Asp486Asp | . | . | . | 6.32E-04 | 4 |
| 7:154595645:C:T | synonymous variant | p.Tyr493Tyr | . | . | . | 7.78E-04 | 9 |
| 7:154596641:C:T | missense variant | p.Thr505Met | . | 34 | 1.65E-05 | 4.86E-05 | 0 |
| 7:154596656:G:A | missense variant | p.Arg510Gln | . | 22.6 | 2.48E-05 | 9.73E-05 | 1 |
| 7:154596668:T:C | missense variant | p.Leu514Pro | . | 25.5 | . | 4.86E-05 | 1 |
| 7:154596669:C:G | synonymous variant | p.Leu514Leu | . | . | . | 4.86E-05 | 0 |
| 7:154598712:C:T | missense variant | p.Thr519Met | . | 26.7 | 1.65E-05 | 9.81E-05 | 0 |
| 7:154598764:C:T | synonymous variant | p.Cys536Cys | . | . | . | 1.96E-04 | 2 |
| 7:154598777:G:A | missense variant | p.Ala541Thr | . | 26.6 | 1.74E-04 | 4.90E-04 | 5 |
| 7:154598789:C:T | missense variant | p.His545Tyr | . | 23.1 | . | 4.91E-05 | 0 |
| 7:154598795:A:G | missense variant | p.Met547Val | . | 0.005 | . | 4.92E-05 | 1 |
| 7:154598806:C:T | synonymous variant | p.Phe550Phe | . | . | . | 4.94E-05 | 1 |
| 7:154598812:C:T | synonymous variant | p.Leu552Leu | . | . | . | 4.95E-05 | 1 |
| 7:154598818:C:T | synonymous variant | p.Cys554Cys | . | . | . | 4.97E-05 | 0 |
| 7:154645486:A:G | splice region variant&intron variant | . | . | . | . | 4.86E-05 | 0 |
| 7:154645517:A:G | missense variant | p.His565Arg | . | 15.82 | . | 4.86E-05 | 1 |
| ***7:154645533:G:T*** | ***missense variant*** | ***p.Lys570Asn*** | ***CO*** | ***11.6*** | ***.*** | ***4.86E-05*** | ***1*** |
| ***7:154645534:A:C*** | ***missense variant*** | ***p.Lys571Gln*** | ***CA*** | ***12.08*** | ***1.04E-03*** | ***7.78E-04*** | ***9*** |
| 7:154645540:A:G | splice region variant&intron variant | . | . | . | . | 4.86E-05 | 1 |
| 7:154659696:C:G | intron variant | . | . | . | . | 4.87E-05 | 1 |
| 7:154659750:A:T | missense variant | p.Asn587Ile | . | 10.61 | 8.28E-06 | 4.86E-05 | 0 |

| | | | | | | | |
|---|---|---|---|---|---|---|---|
| 7:154659756:G:A | missense variant | p.Arg589Gln | . | 17.04 | 5.79E-05 | 4.86E-05 | 0 |
| 7:154659766:T:C | synonymous variant | p.Pro592Pro | . | . | . | 2.43E-04 | 2 |
| 7:154659784:C:T | synonymous variant | p.Asp598Asp | . | . | . | 4.86E-05 | 1 |
| 7:154659792:T:C | missense variant | p.Ile601Thr | . | 7.708 | 5.80E-05 | 4.86E-05 | 1 |
| 7:154664371:C:T | synonymous variant | p.Thr617Thr | . | . | . | 4.89E-05 | 0 |
| 7:154664404:G:A | splice donor variant&intron variant | . | . | 26.8 | . | 4.89E-05 | 1 |
| 7:154667638:A:T | missense variant | p.Ser636Cys | . | 22.5 | . | 4.88E-05 | 1 |
| 7:154667643:G:A | synonymous variant | p.Val637Val | . | . | . | 3.50E-01 | 697/2460/2419 |
| 7:154667673:G:A | synonymous variant | p.Thr647Thr | . | . | . | 4.88E-05 | 0 |
| 7:154667688:C:T | synonymous variant | p.Ser652Ser | . | . | . | 4.89E-05 | 1 |
| 7:154667691:C:T | synonymous variant | p.His653His | . | . | . | 4.89E-05 | 0 |
| 7:154667692:G:A | missense variant | p.Gly654Ser | . | 20.7 | 2.16E-03 | 9.77E-05 | 1 |
| ***7:154667695:G:A*** | ***missense variant*** | ***p.Ala655Thr*** | ***CA*** | ***16.64*** | ***1.65E-05*** | ***4.89E-05*** | ***1*** |
| 7:154667715:C:T | synonymous variant | p.Asp661Asp | . | . | . | 4.90E-05 | 1 |
| 7:154667716:G:A | missense variant | p.Gly662Ser | . | 29.8 | 2.48E-05 | 4.90E-05 | 0 |
| 7:154667727:C:T | synonymous variant | p.Ser665Ser | . | . | . | 4.92E-05 | 1 |
| 7:154667728:G:A | missense variant | p.Gly666Ser | . | 26.2 | . | 4.92E-05 | 0 |
| 7:154672631:G:A | synonymous variant | p.Thr704Thr | . | . | . | 4.86E-05 | 1 |
| 7:154672632:C:T | missense variant | p.Arg705Cys | . | 26.4 | 8.26E-06 | 4.87E-05 | 0 |
| 7:154672634:C:T | synonymous variant | p.Arg705Arg | . | . | . | 4.87E-05 | 1 |
| 7:154672646:T:C | synonymous variant | p.Phe709Phe | . | . | . | 1.83E-01 | 198/1700/3749 |
| 7:154672652:G:A | splice region variant&synonymous variant | p.Lys711Lys | . | . | . | 4.87E-05 | 0 |
| 7:154677402:G:A | synonymous variant | p.Gln731Gln | . | . | . | 1.47E-04 | 3 |
| 7:154677414:C:T | synonymous variant | p.Cys735Cys | . | . | . | 3.43E-03 | 39 |
| 7:154677462:G:A | splice region variant&intron variant | . | . | . | . | 1.01E-04 | 0 |
| 7:154679392:C:T | missense variant | p.Ala751Val | . | 24.1 | 8.26E-06 | 9.73E-05 | 0 |
| 7:154679447:G:A | splice region variant&intron variant | . | . | . | . | 3.84E-02 | 9/430/5240 |
| 7:154680976:G:T | splice region variant&intron variant | . | . | . | . | 4.86E-05 | 1 |
| 7:154680993:A:T | synonymous variant | p.Val772Val | . | . | . | 9.73E-05 | 2 |
| 7:154680994:G:T | missense variant | p.Ala773Ser | . | 22.5 | . | 4.86E-05 | 1 |
| 7:154681001:G:A | missense variant | p.Arg775Gln | . | 27.5 | 2.48E-05 | 4.86E-05 | 1 |
| 7:154681008:C:T | synonymous variant | p.Ser777Ser | . | . | . | 5.84E-04 | 6 |
| ***7:154681009:G:A*** | ***missense variant*** | ***p.Ala778Thr*** | ***CO*** | ***15.25*** | ***1.47E-03*** | ***2.34E-03*** | ***28*** |
| 7:154681010:C:T | missense variant | p.Ala778Val | . | 17.44 | 2.56E-04 | 9.73E-05 | 2 |

| | | | | | | | |
|---|---|---|---|---|---|---|---|
| 7:154681011:G:A | synonymous variant | p.Ala778Ala | . | . | . | 9.73E-05 | 1 |
| 7:154681014:G:A | synonymous variant | p.Leu779Leu | . | . | . | 4.86E-05 | 1 |
| 7:154681048:G:A | missense variant | p.Ala791Thr | . | 24.6 | . | 4.86E-05 | 0 |
| 7:154681050:C:T | synonymous variant | p.Ala791Ala | . | . | . | 1.18E-01 | 77/1198/4404 |
| 7:154681174:T:A | synonymous variant | p.Ile795Ile | . | . | . | 4.87E-05 | 0 |
| 7:154681203:C:A | missense variant | p.Thr805Lys | . | 15.49 | . | 4.86E-05 | 0 |
| 7:154681216:G:A | synonymous variant | p.Arg809Arg | . | . | . | 1.43E-01 | 113/1430/4135 |
| 7:154681220:A:G | missense variant | p.Lys811Glu | . | 15.52 | 2.15E-04 | 3.89E-04 | 3 |
| 7:154684041:C:T | splice region variant&intron variant | . | . | . | . | 4.86E-05 | 0 |
| 7:154684068:T:C | missense variant | p.Phe826Leu | . | 2.809 | 8.26E-06 | 4.86E-05 | 1 |
| 7:154684078:C:A | missense variant | p.Ser829Tyr | . | 13.41 | . | 4.86E-05 | 1 |
| 7:154684082:C:T | synonymous variant | p.Ser830Ser | . | . | . | 9.73E-05 | 1 |
| 7:154684153:T:C | missense variant | p.Leu854Pro | . | 5.639 | 3.21E-01 | 2.74E-01 | 439/2277/2962 |

* SNP-ID is in GRCh37 reference

** MAF as in ExAC non-Finnish European population

In italics and bold are highlighted previously reported variants in DPP6 by Cacace et al.

| ADSP | | | EOAD | | | | | FASe | | | | |
|---|---|---|---|---|---|---|---|---|---|---|---|---|
| Controls (n=4601) | OR | P | MAF | Cases (n=1383) | Controls (n=3684) | OR | P | MAF | Cases (n=5656) | Controls (n=4601) | OR | P |
| 1 | 0 | 0.266 | . | . | . | . | . | . | . | . | . | . |
| 1 | 0 | 0.265 | . | . | . | . | . | . | . | . | . | . |
| . | . | . | . | . | . | . | . | 2.91E-04 | 1 | 0 | NA | 0.596 |
| 0 | NA | 0.368 | . | . | . | . | . | . | . | . | . | . |
| 0 | NA | 0.368 | . | . | . | . | . | . | . | . | . | . |
| 0 | NA | 0.368 | . | . | . | . | . | . | . | . | . | . |
| 1 | 0.81 | 0.881 | . | . | . | . | . | . | . | . | . | . |
| 1 | 0.81 | 0.882 | . | . | . | . | . | . | . | . | . | . |
| 9 | 0.90 | 0.818 | 1.53E-03 | 5 | 11 | 1.27 | 0.659 | . | . | . | . | . |
| 1 | 0 | 0.266 | . | . | . | . | . | . | . | . | . | . |
| 0 | NA | 0.368 | . | . | . | . | . | . | . | . | . | . |
| 0 | NA | 0.368 | . | . | . | . | . | . | . | . | . | . |
| 0 | NA | 0.368 | . | . | . | . | . | . | . | . | . | . |
| 4 | 0.81 | 0.765 | . | . | . | . | . | . | . | . | . | . |
| 0 | NA | 0.368 | . | . | . | . | . | . | . | . | . | . |
| 1 | 0 | 0.266 | . | . | . | . | . | . | . | . | . | . |
| 1 | 0 | 0.267 | . | . | . | . | . | . | . | . | . | . |
| 1 | 0 | 0.267 | . | . | . | . | . | . | . | . | . | . |
| 3 | 0 | 0.054 | . | . | . | . | . | . | . | . | . | . |
| 1 | 0.81 | 0.881 | . | . | . | . | . | . | . | . | . | . |
| 1 | 0 | 0.267 | . | . | . | . | . | . | . | . | . | . |
| 51/760/3786 | 1.05 | 0.279 | . | . | . | . | . | . | . | . | . | . |
| 136 | 1.08 | 0.504 | . | . | . | . | . | . | . | . | . | . |
| 0 | NA | 0.368 | . | . | . | . | . | . | . | . | . | . |
| 1 | 0 | 0.267 | . | . | . | . | . | . | . | . | . | . |
| 0 | NA | 0.368 | . | . | . | . | . | . | . | . | . | . |
| 2 | 0.41 | 0.446 | . | . | . | . | . | . | . | . | . | . |
| 1 | 0 | 0.267 | . | . | . | . | . | . | . | . | . | . |
| 3 | 0 | 0.054 | . | . | . | . | . | . | . | . | . | . |
| 1 | 0.81 | 0.882 | . | . | . | . | . | . | . | . | . | . |

| | | | | | | | | | | | | |
|---|---|---|---|---|---|---|---|---|---|---|---|---|
| 0 | NA | 0.367 | . | . | . | . | . | . | . | . | . | . |
| 52 | 1.05 | 0.809 | . | . | . | . | . | . | . | . | . | . |
| 46/611/3726 | 0.97 | 0.596 | . | . | . | . | . | . | . | . | . | . |
| 138 | 1.05 | 0.650 | . | . | . | . | . | . | . | . | . | . |
| 1 | 0 | 0.268 | . | . | . | . | . | . | . | . | . | . |
| 0 | NA | 0.201 | . | . | . | . | . | . | . | . | . | . |
| 382/1857/2357 | 1.02 | 0.616 | . | . | . | . | . | . | . | . | . | . |
| 45 | 1.21 | 0.329 | . | . | . | . | . | . | . | . | . | . |
| 0 | NA | 0.368 | . | . | . | . | . | . | . | . | . | . |
| ***1*** | ***2.43*** | ***0.427*** | . | . | . | . | . | . | . | . | . | . |
| 0 | NA | 0.368 | . | . | . | . | . | . | . | . | . | . |
| 44/793/3764 | 1.02 | 0.740 | . | . | . | . | . | . | . | . | . | . |
| . | . | . | . | . | . | . | . | 8.74E-04 | 3 | 0 | NA | 0.358 |
| 0 | NA | 0.368 | 9.53E-05 | 1 | 0 | NA | 0.095 | . | . | . | . | . |
| 0 | NA | 0.368 | . | . | . | . | . | . | . | . | . | . |
| 1 | 0 | 0.267 | . | . | . | . | . | . | . | . | . | . |
| 5 | 1.30 | 0.648 | . | . | . | . | . | . | . | . | . | . |
| ***1*** | ***0*** | ***0.267*** | . | . | . | . | . | . | . | . | . | . |
| 0 | NA | 0.368 | . | . | . | . | . | . | . | . | . | . |
| 1 | 0 | 0.267 | . | . | . | . | . | . | . | . | . | . |
| 1 | 0 | 0.267 | . | . | . | . | . | . | . | . | . | . |
| 0 | NA | 0.368 | . | . | . | . | . | . | . | . | . | . |
| 1 | 2.43 | 0.427 | . | . | . | . | . | . | . | . | . | . |
| 1 | 0.81 | 0.882 | . | . | . | . | . | . | . | . | . | . |
| 0 | NA | 0.368 | . | . | . | . | . | . | . | . | . | . |
| 0 | NA | 0.368 | . | . | . | . | . | . | . | . | . | . |
| 45/706/3850 | 0.98 | 0.700 | . | . | . | . | . | . | . | . | . | . |
| ***1*** | ***0.81*** | ***0.882*** | . | . | . | . | . | . | . | . | . | . |
| 1 | 0 | 0.267 | . | . | . | . | . | . | . | . | . | . |
| 0 | NA | 0.368 | . | . | . | . | . | . | . | . | . | . |
| 1 | 0 | 0.266 | . | . | . | . | . | . | . | . | . | . |
| 0 | NA | 0.368 | . | . | . | . | . | . | . | . | . | . |
| 0 | NA | 0.368 | . | . | . | . | . | . | . | . | . | . |
| ***0*** | ***NA*** | ***0.203*** | . | . | . | . | . | . | . | . | . | . |

| | | | | | | | | | | | | |
|---|---|---|---|---|---|---|---|---|---|---|---|---|
| 1 | 0 | 0.267 | . | . | . | . | . | . | . | . | . | . |
| 0 | NA | 0.368 | . | . | . | . | . | . | . | . | . | . |
| 0 | NA | 0.369 | . | . | . | . | . | . | . | . | . | . |
| 1 | 0 | 0.267 | . | . | . | . | . | . | . | . | . | . |
| 0 | NA | 0.368 | . | . | . | . | . | . | . | . | . | . |
| 1 | 0 | 0.267 | . | . | . | . | . | . | . | . | . | . |
| 0 | NA | 0.368 | . | . | . | . | . | . | . | . | . | . |
| 0 | NA | 0.369 | . | . | . | . | . | . | . | . | . | . |
| 1 | 0 | 0.267 | . | . | . | . | . | . | . | . | . | . |
| 1 | 0 | 0.267 | . | . | . | . | . | . | . | . | . | . |
| 0 | NA | 0.368 | . | . | . | . | . | . | . | . | . | . |
| 3 | 0 | 0.054 | . | . | . | . | . | . | . | . | . | . |
| 0 | NA | 0.368 | . | . | . | . | . | . | . | . | . | . |
| 9 | 0.36 | 0.076 | . | . | . | . | . | . | . | . | . | . |
| 7 | 1.04 | 0.935 | . | . | . | . | . | . | . | . | . | . |
| 1 | 0 | 0.267 | . | . | . | . | . | . | . | . | . | . |
| 1 | 0.81 | 0.881 | . | . | . | . | . | . | . | . | . | . |
| 0 | NA | 0.368 | . | . | . | . | . | . | . | . | . | . |
| 1 | 0 | 0.267 | . | . | . | . | . | . | . | . | . | . |
| 2 | 0 | 0.117 | . | . | . | . | . | . | . | . | . | . |
| 2 | 0.81 | 0.834 | . | . | . | . | . | . | . | . | . | . |
| 5 | 0.81 | 0.742 | . | . | . | . | . | . | . | . | . | . |
| 1 | 0 | 0.267 | . | . | . | . | . | . | . | . | . | . |
| 0 | NA | 0.367 | . | . | . | . | . | . | . | . | . | . |
| 0 | NA | 0.368 | . | . | . | . | . | . | . | . | . | . |
| 0 | NA | 0.368 | . | . | . | . | . | . | . | . | . | . |
| 1 | 0 | 0.267 | . | . | . | . | . | . | . | . | . | . |
| 1 | 0 | 0.267 | . | . | . | . | . | . | . | . | . | . |
| 0 | NA | 0.368 | . | . | . | . | . | . | . | . | . | . |
| ***0*** | ***NA*** | ***0.368*** | . | . | . | . | . | . | . | . | . | . |
| ***7*** | ***1.04*** | ***0.935*** | . | . | . | . | . | . | . | . | . | . |
| 0 | NA | 0.368 | . | . | . | . | . | . | . | . | . | . |
| 0 | NA | 0.368 | . | . | . | . | . | . | . | . | . | . |
| 1 | 0 | 0.267 | . | . | . | . | . | . | . | . | . | . |

| | | | | | | | | | | | | |
|---|---|---|---|---|---|---|---|---|---|---|---|---|
| 1 | 0 | 0.267 | . | . | . | . | . | . | . | . | . | . |
| 3 | 0.54 | 0.493 | . | . | . | . | . | . | . | . | . | . |
| 0 | NA | 0.368 | . | . | . | . | . | . | . | . | . | . |
| 0 | NA | 0.368 | . | . | . | . | . | . | . | . | . | . |
| 1 | 0 | 0.266 | . | . | . | . | . | . | . | . | . | . |
| 0 | NA | 0.369 | . | . | . | . | . | . | . | . | . | . |
| 0 | NA | 0.369 | 9.56E-05 | 1 | 0 | NA | 0.096 | . | . | . | . | . |
| 574/1999/1863 | 0.96 | 0.179 | | . | . | . | . | . | . | . | . | . |
| 1 | 0 | 0.266 | 1.91E-04 | 1 | 1 | 2.78 | 0.451 | . | . | . | . | . |
| 0 | NA | 0.369 | . | . | . | . | . | . | . | . | . | . |
| 1 | 0 | 0.266 | . | . | . | . | . | . | . | . | . | . |
| 1 | 0.81 | 0.879 | . | . | . | . | . | . | . | . | . | . |
| ***0*** | ***NA*** | ***0.369*** | ***1.91E-04*** | ***1*** | ***1*** | ***2.78*** | ***0.451*** | ***.*** | ***.*** | ***.*** | ***.*** | ***.*** |
| 0 | NA | 0.369 | . | . | . | . | . | . | . | . | . | . |
| 1 | 0 | 0.266 | . | . | . | . | . | . | . | . | . | . |
| 0 | NA | 0.370 | . | . | . | . | . | . | . | . | . | . |
| 1 | 0 | 0.265 | . | . | . | . | . | . | . | . | . | . |
| 0 | NA | 0.368 | . | . | . | . | . | . | . | . | . | . |
| 1 | 0 | 0.267 | . | . | . | . | . | . | . | . | . | . |
| 0 | NA | 0.368 | . | . | . | . | . | . | . | . | . | . |
| 165/1302/3088 | 1.04 | 0.237 | . | . | . | . | . | . | . | . | . | . |
| 1 | 0 | 0.266 | . | . | . | . | . | . | . | . | . | . |
| 0 | NA | 0.118 | 9.56E-05 | 1 | 0 | NA | 0.095 | . | . | . | . | . |
| 31 | 1.03 | 0.915 | 3.06E-03 | 8 | 24 | 0.93 | 0.852 | 4.95E-03 | 12 | 3 | 1.13 | 0.854 |
| 2 | 0 | 0.118 | . | . | . | . | . | . | . | . | . | . |
| 2 | 0 | 0.116 | . | . | . | . | . | . | . | . | . | . |
| 5/332/4263 | 1.06 | 0.400 | . | . | . | . | . | . | . | . | . | . |
| 0 | NA | 0.368 | . | . | . | . | . | . | . | . | . | . |
| 0 | NA | 0.203 | . | . | . | . | . | . | . | . | . | . |
| 0 | NA | 0.368 | . | . | . | . | . | . | . | . | . | . |
| 0 | NA | 0.368 | . | . | . | . | . | . | . | . | . | . |
| 6 | 0.81 | 0.715 | . | . | . | . | . | . | . | . | . | . |
| ***20*** | ***1.14*** | ***0.666*** | . | . | . | . | . | . | . | . | . | . |
| 0 | NA | 0.203 | . | . | . | . | . | . | . | . | . | . |

| | | | | | | | | | | | | |
|---|---|---|---|---|---|---|---|---|---|---|---|---|
| 1 | 0.81 | 0.882 | . | . | . | . | . | . | . | . | . | . |
| 0 | NA | 0.368 | . | . | . | . | . | . | . | . | . | . |
| 1 | 0 | 0.267 | . | . | . | . | . | . | . | . | . | . |
| 78/926/3597 | 1.01 | 0.749 | . | . | . | . | . | . | . | . | . | . |
| 1 | 0 | 0.267 | . | . | . | . | . | . | . | . | . | . |
| 1 | 0 | 0.267 | . | . | . | . | . | . | . | . | . | . |
| 99/1093/3406 | 1.05 | 0.269 | . | . | . | . | . | . | . | . | . | . |
| 5 | 0.49 | 0.313 | . | . | . | . | . | . | . | . | . | . |
| 1 | 0 | 0.267 | . | . | . | . | . | . | . | . | . | . |
| 0 | NA | 0.368 | . | . | . | . | . | . | . | . | . | . |
| 0 | NA | 0.368 | . | . | . | . | . | . | . | . | . | . |
| 0 | NA | 0.203 | . | . | . | . | . | . | . | . | . | . |
| 352/1782/2465 | 1.04 | 0.228 | 0.27 | 667 | 1812 | 1.03 | 0.507 | . | . | . | . | . |
