## Supplementary-table3 for "DPP6 gene in European American Alzheimer’s Disease": 2020-10-DPP6-Sup-table-3.pdf

**Supplementary Table 3. V**ariants included in the CADD>20 gene-based analyses in each of the cohorts investigated: ADSP, EOAD and FASe.

| SNP-ID* | EFFECT | p.CHAN GE | direction in Cacace | CADD | MAF-ExAC** | MAF | Cases (n=5656) |
|---|---|---|---|---|---|---|---|
| 7:154667692:G:A | missense variant | p.Gly654Ser | . | 20.7 | 2.16E-03 | 9.77E-05 | 1 |
| 7:154429580:A:G | missense variant | p.His226Arg | . | 21.2 | . | 4.87E-05 | 1 |
| ***7:154564586:A:G*** | ***missense variant*** | ***p.His357Arg*** | ***CA*** | ***21.7*** | ***.*** | ***9.73E-05*** | ***2*** |
| ***7:154461074:C:A*** | ***missense variant*** | ***p.Pro229Thr*** | ***CA*** | ***22.1*** | ***.*** | ***1.95E-04*** | ***3*** |
| 7:154595580:A:G | missense variant | p.Ser472Gly | . | 22.3 | . | 4.86E-05 | 1 |
| 7:154667638:A:T | missense variant | p.Ser636Cys | . | 22.5 | . | 4.88E-05 | 1 |
| 7:154680994:G:T | missense variant | p.Ala773Ser | . | 22.5 | . | 4.86E-05 | 1 |
| 7:154596656:G:A | missense variant | p.Arg510Gln | . | 22.6 | 2.48E-05 | 9.73E-05 | 1 |
| 7:154263951:G:A | missense variant | p.Glu193Lys | . | 22.8 | 2.48E-05 | 4.87E-05 | 1 |
| 7:154593078:T:C | missense variant | p.Val438Ala | . | 22.9 | . | 4.86E-05 | 0 |
| 7:154593156:C:T | missense variant | p.Thr464Met | . | 23 | . | 4.88E-05 | 1 |
| 7:154598789:C:T | missense variant | p.His545Tyr | . | 23.1 | . | 4.91E-05 | 0 |
| 7:154461105:A:C | missense variant | p.Asn239Thr | . | 23.4 | . | 4.86E-05 | 1 |
| 7:154519513:C:T | missense variant | p.His267Tyr | . | 23.8 | 3.31E-05 | 4.86E-05 | 0 |
| 7:154679392:C:T | missense variant | p.Ala751Val | . | 24.1 | 8.26E-06 | 9.73E-05 | 0 |
| 7:154681048:G:A | missense variant | p.Ala791Thr | . | 24.6 | . | 4.86E-05 | 0 |
| 7:154561240:G:A | missense variant | p.Gly333Ser | . | 25 | 1.66E-04 | 4.87E-05 | 1 |
| 7:154587577:C:T | missense variant | p.Ala428Val | . | 25.1 | . | 4.86E-05 | 1 |
| 7:154587563:G:T | missense variant | p.Glu423Asp | . | 25.2 | . | 4.86E-05 | 0 |
| 7:154596668:T:C | missense variant | p.Leu514Pro | . | 25.5 | . | 4.86E-05 | 1 |
| 7:154263988:A:G | missense variant | p.Tyr205Cys | . | 25.8 | . | 4.87E-05 | 0 |
| ***7:154561208:G:A*** | ***missense variant*** | ***p.Arg322His*** | ***CA*** | ***25.8*** | ***1.08E-04*** | ***9.73E-05*** | ***1*** |
| 7:154564555:G:T | missense variant&splice region variant | p.Ala347Ser | . | 26 | 8.27E-06 | 4.86E-05 | 1 |
| 7:154667728:G:A | missense variant | p.Gly666Ser | . | 26.2 | . | 4.92E-05 | 0 |
| 7:154672632:C:T | missense variant | p.Arg705Cys | . | 26.4 | 8.26E-06 | 4.87E-05 | 0 |
| 7:154598777:G:A | missense variant | p.Ala541Thr | . | 26.6 | 1.74E-04 | 4.90E-04 | 5 |
| 7:154598712:C:T | missense variant | p.Thr519Met | . | 26.7 | 1.65E-05 | 9.81E-05 | 0 |
| 7:154561183:G:A | missense variant | p.Ala314Thr | . | 27.1 | 3.31E-05 | 4.86E-05 | 1 |
| 7:154461135:C:T | missense variant | p.Pro249Leu | . | 27.3 | . | 4.86E-05 | 1 |
| 7:154681001:G:A | missense variant | p.Arg775Gln | . | 27.5 | 2.48E-05 | 4.86E-05 | 1 |

| | | | | | | | |
|---|---|---|---|---|---|---|---|
| 7:154564646:G:T | missense variant | p.Arg377Leu | . | 27.6 | . | 4.88E-05 | 1 |
| 7:154561276:C:T | missense variant | p.Pro345Ser | . | 28.5 | 8.27E-06 | 4.94E-05 | 1 |
| 7:154667716:G:A | missense variant | p.Gly662Ser | . | 29.8 | 2.48E-05 | 4.90E-05 | 0 |
| ***7:154519535:G:A*** | ***missense variant*** | ***p.Arg274His*** | ***CA*** | ***34*** | ***8.27E-06*** | ***4.86E-05*** | ***0*** |
| 7:154593084:C:T | missense variant | p.Ser440Phe | . | 34 | . | 4.86E-05 | 0 |
| 7:154596641:C:T | missense variant | p.Thr505Met | . | 34 | 1.65E-05 | 4.86E-05 | 0 |
| 7:154587586:AC:A | frameshift variant | p.His431fs | . | . | . | 4.87E-05 | 1 |

* SNP-ID is in GRCh37 reference

** MAF as in ExAC non-Finnish European population

In italics and bold are highlighted previously reported variants in DPP6 by Cacace et al.

| ADSP | | | EOAD | | | | | FASe | | | | |
|---|---|---|---|---|---|---|---|---|---|---|---|---|
| Controls (n=4601) | OR | P | MAF | Cases (n=1383) | Controls (n=3684) | OR | P | MAF | Cases (n=5656) | Controls (n=4601) | OR | P |
| 1 | 0.81 | 0.879 | . | . | . | . | . | . | . | . | . | . |
| 0 | NA | 0.368 | . | . | . | . | . | . | . | . | . | . |
| ***0*** | ***NA*** | ***0.203*** | . | . | . | . | . | . | . | . | . | . |
| ***1*** | ***2.43*** | ***0.427*** | . | . | . | . | . | . | . | . | . | . |
| 0 | NA | 0.368 | . | . | . | . | . | . | . | . | . | . |
| 0 | NA | 0.369 | 9.56E-05 | 1 | 0 | NA | 0.096 | . | . | . | . | . |
| 0 | NA | 0.368 | . | . | . | . | . | . | . | . | . | . |
| 1 | 0.81 | 0.881 | . | . | . | . | . | . | . | . | . | . |
| 0 | NA | 0.368 | . | . | . | . | . | . | . | . | . | . |
| 1 | 0.00 | 0.267 | . | . | . | . | . | . | . | . | . | . |
| 0 | NA | 0.368 | . | . | . | . | . | . | . | . | . | . |
| 1 | 0.00 | 0.267 | . | . | . | . | . | . | . | . | . | . |
| 0 | NA | 0.368 | . | . | . | . | . | . | . | . | . | . |
| 1 | 0.00 | 0.267 | . | . | . | . | . | . | . | . | . | . |
| 2 | 0.00 | 0.116 | . | . | . | . | . | . | . | . | . | . |
| 1 | 0.00 | 0.267 | . | . | . | . | . | . | . | . | . | . |
| 0 | NA | 0.368 | . | . | . | . | . | . | . | . | . | . |
| 0 | NA | 0.368 | . | . | . | . | . | . | . | . | . | . |
| 1 | 0.00 | 0.267 | . | . | . | . | . | . | . | . | . | . |
| 0 | NA | 0.368 | . | . | . | . | . | . | . | . | . | . |
| 1 | 0.00 | 0.266 | . | . | . | . | . | . | . | . | . | . |
| ***1*** | ***0.81*** | ***0.882*** | . | . | . | . | . | . | . | . | . | . |
| 0 | NA | 0.368 | . | . | . | . | . | . | . | . | . | . |
| 1 | 0.00 | 0.265 | . | . | . | . | . | . | . | . | . | . |
| 1 | 0.00 | 0.267 | . | . | . | . | . | . | . | . | . | . |
| 5 | 0.81 | 0.742 | . | . | . | . | . | . | . | . | . | . |
| 2 | 0.00 | 0.117 | . | . | . | . | . | . | . | . | . | . |
| 0 | NA | 0.368 | . | . | . | . | . | . | . | . | . | . |
| 0 | NA | 0.368 | 9.53E-05 | 1 | 0 | NA | 0.095 | . | . | . | . | . |
| 0 | NA | 0.368 | . | . | . | . | . | . | . | . | . | . |

| | | | | | | | | | | | | |
|---|---|---|---|---|---|---|---|---|---|---|---|---|
| 0 | NA | 0.369 | . | . | . | . | . | . | . | . | . | . |
| 0 | NA | 0.368 | . | . | . | . | . | . | . | . | . | . |
| 1 | 0.00 | 0.266 | . | . | . | . | . | . | . | . | . | . |
| ***1*** | ***0.00*** | ***0.267*** | . | . | . | . | . | . | . | . | . | . |
| 1 | 0.00 | 0.267 | . | . | . | . | . | . | . | . | . | . |
| 1 | 0.00 | 0.267 | . | . | . | . | . | . | . | . | . | . |
| 0 | NA | 0.369 | . | . | . | . | . | . | . | . | . | . |
